# From Classroom to Clinical: A Mixed methods study of the Impact of Simulation Based Education on Final Year Physiotherapy Students

**DOI:** 10.64898/2026.08.21.26360984

**Authors:** Ciara McGurn, Leslie George

## Abstract

**Objectives:** This study aimed to examine physiotherapy student’s reaction and learning, following Simulation Based Education (SBE) during a Cardiorespiratory module. A further aim was to see if any learning translated into clinical placement.

**Design:** A mixed methods research design consisting of a questionnaire (Phase1) after the activity which was underpinned by the Kirkpatrick model of evaluation. This was followed by a focus groups (Phase 2) after completion of clinical placement.

**Participants:** 92 final year physiotherapy students at a single institution were eligible to participate in the SBE session with n=81 (88%) students completing the survey, and 8 students participating across 2 focus group sessions.

**Results:** Survey: Over 80% of students strongly agreed on a positive initial reaction to SBE. Learning yielded a 76% and above response of strongly agree in all areas except confidence. Students valued SBE as a preferred learning and teaching strategy and wanted more. They welcomed SBE as a supplement but not substitute for clinical placement. Students felt skills learned could be transferred into all areas of clinical practice,, namely communication and decision making.

**Conclusion:** Students rated SBE positively with development of transferable non-technical skills. Reaction to SBE was high in terms of relevance, engagement and satisfaction. Self-perceived confidence, although positive, was the comparatively lowest scoring of the domains. Students advocated SBE as a supplement rather than a substitute for clinical placement preferring a hybrid approach.

**Contribution of the Paper:**

- Adds to the positive body of evidence which already exists towards SBE, especially in the development of non-technical skills.
- Suggestions are made that SBE can’t make students feel fully confident in preparation for clinical placement.
- While students in this study expressed that they wanted more SBE and earlier, this paper found that students would not advocate SBE replacing clinical placement.

## INTRODUCTION

A consensus definition for Simulation Based Education (SBE) describes SBE as a ‘*tool that supports development through experiential learning by replicating conditions that resemble real life.’* [1] The definition goes further to highlight the importance of psychological safety during SBE, so participants can learn from their mistakes. This is in keeping with the notion of making mistakes while learning, without exposing patients to the associated risks [2]. SBE strives to go beyond theory and clinical skills and develop non-technical skills through the understanding of roles and responsibilities, promoting teamwork and communication, thus improving patient outcomes [3]. The scope of SBE is rapidly developing beyond role play and manikins to include immersive environments, virtual reality and artificial intelligence [4]. Despite technological advances, learning theory has not changed rapidly [5]. For learning to occur, the core elements of SBE remain; namely pre-brief, the scenario itself and the essential debrief (reflection) [6]. The Adoption of SBE as an educational tool in physiotherapy was recommended by the KNOWBEST report [7].

The Kirkpatrick Framework is a widely used model that evaluates training programmes and transfer of learning outcomes [8]. This self-reported model consists of 4 levels: Reaction, Learning, Behaviour and Results, and adopts a systematic approach which delves further within each level to ascertain the rich layers of learning. Within physiotherapy, a recent scoping review acknowledged the increased output in terms of SBE evidence over the last 10 years [9]. This review observed however that much of this research is focused on learner satisfaction (equating to Kirkpatrick Levels 1 and 2), with a paucity of evidence exploring the impact of translation of learning into clinical practice. The lack of evidence to support translation into clinical practice is supported by other sources [10,11]. SBE may assist in better preparing students for successful integration into the dynamic healthcare environment however future research should explore the long-term retention and application of knowledge acquired through SBE in real clinical scenarios [12]. Further research is therefore required to understand if any benefits gained in simulation are translated into the clinical environment eliciting behavioural change i.e. Kirkpatrick Level 3.

The aim of this study was to ascertain initial physiotherapy student reaction and learning following a SBE session then to further explore transfer of learning into clinical placement.

## METHODS

### Design

A sequential explanatory mixed methods research design was adopted. This study comprised of two distinct but related phases comprising quantitative data collection via a survey and qualitative data via two focus group sessions. Ethical approval was granted by a University Nursing and Health Sciences Filter committee (Reference: FCNUR-24-072). Data collection took place between October 2024 and January 2025. Electronic consent forms were completed by participants before each phase of the study.

### Participants

Final year undergraduate physiotherapy students (n=92) enrolled at a single institution were approached to participate in the study. As part of their educational programme all enrolled students undertook a SBE session in Semester 1. The students were e-mailed by the Chief Investigator (CI) two weeks prior to the SBE session, inviting them to participate in the research. To be eligible to participate students were required to be registered in a cardiorespiratory module and available to attend the SBE session delivered as part of this module. Students of all genders, over 18 years of age were included, and students had to provide informed consent. Anyone not meeting these criteria were excluded. As part of the survey, those participants who wished to be approached to take part in the focus group research were asked to supply their e mail address. Students who supplied their contact details were then contacted via e mail to indicate their availability for the focus group sessions. To be included in the focus groups participants needed to have completed the survey, completed a placement after the SBE session and provided informed consent. Participants were excluded if they were unable to complete placement for medical or any other reasons and therefore didn’t meet the placement learning outcomes. Participants who did not complete Phase 1 were also excluded. Participation was voluntary and did not impact on student’s educational experience/ outcome.

### Interventions

As part of the curriculum design all final year physiotherapy students undertake a cardiorespiratory module which includes a SBE session. Students participated in 3 scenarios each lasting approximately 40 minutes (5 minutes pre-brief, 10 minutes for the scenario with a 25 minutes debrief). The scenarios covered the following topics:

1. Identification and management of a partially obstructed airway
2. Cardiac arrest situation with a tracheostomy patient
3. Cardiovascular/ Respiratory instability while ‘bagging’ a ventilated patient

Phase 1 of the study took place in a clinical skills suite and phase 2 focus groups took place online via Microsoft Teams.

#### Phase 1: Survey

Following the SBE session students completed an electronic survey, emailed to students and completed via the JISC platform.

#### Phase 2: Focus Groups

A semi structured interview was undertaken around 4 themes – Overall Impression, Learning, Impact/Transferability and Future. Questions were developed by the investigators to address the study aims and were mostly open ended to encourage discussion. Questions were designed based on Level 3 of the Kirkpatrick model of evaluation, to gauge behavioural change. Eighteen potential questions were used to guide the focus groups, however the aim was to guide the conversation and allow relevant tangential discussions to occur. Flexibility allowed for unanticipated themes to emerge thereby gleaning further qualitative information.

The focus groups were held online via Microsoft Teams and recorded with permission from participants. The focus groups were held after a clinical placement in order to maximise the relevance of the topics being explored

### Outcomes

The survey was created by the investigators (following the Kirkpatrick framework) to measure Levels 1(Reaction) and 2 (Learning). Questions in each section were generated from examples provided by the Kirkpatrick website (The Kirkpatrick Model (kirkpatrickpartners.com). Questions were also reworked using the Satisfaction with Simulation Experience Scale (SSE) [13]. Level 1(reaction) questioned participants on engagement, relevance, and satisfaction. Level 2 (learning) asked about skills/knowledge, attitude, confidence and commitment. Questions were asked in the positive form, and responses took the form of a 5 point Likert scale (strongly disagree, disagree, neither agree or disagree, agree and strongly agree).

### Statistical Analysis

The frequency of Likert responses was collated for Levels 1 and 2 of the survey, specifically the modal response.

The focus group sessions were transcribed verbatim. The transcript was imported into NVIVO version 14.23.3.61. Thematic analysis was utilised by the authors and data was handled following a six-step guide [14]. The CI led the thematic analysis by first familiarising themselves with the transcript. Adopting an inductive analysis approach, the data was scrutinised line by line and code generated aligned to the objectives of the study. Sixteen codes were identified which were then grouped into themes. Analyst triangulation took place involving regular discussion with the second researcher. Researchers kept a record / audit trail of discussions. All codes were included in analysis to maintain reflexivity.

## RESULTS

The survey generated a response rate of 88% (n=81). From this sample 10% (n=8) participated in the focus groups. Participants were predominantly female and aged 20-25 years. Placement areas for those undertaking the survey varied but half of the participants in the focus group were in a musculoskeletal outpatient setting (Table 1).

**Table 1:** Participant Demographics.

|  | PHASE 1 (SURVEY) | PHASE 2 (FOCUS GROUPS) |
| --- | --- | --- |
| Gender | Female n=56 (69%)<br>Male n=25 (31%) | Female n=7 (88%)<br>Male n=1 (12%) |
| Age | < 20 n=2 (2%)<br>20-25 n=63 (78%)<br>26-30 n=8 (10%)<br>31-35 n=4 (5%)<br>36-40 n=3 (4%)<br>41-45 n=1 (1%) | 20-25 n=6 (75%)<br>31-35 n=1 (13%)<br>41-45 n=1 (13%) |
| Placement Area | Surgical n=1 (1%)<br>Medical n=16 (20%)<br>Critical Care n=4 (5%)<br>MSK n=19 (23%)<br>Orthopaedics n=5 (6%)<br>Neuro n=10 (12%)<br>Paediatrics n=10 (12%)<br>Elderly Care n=3 (4%)<br>Other n=13 (16%) | MSK n=4 (50%)<br>Paediatrics n=2 (25%)<br>Other n=2 (25%) |

The frequency of responses for Level 1 (Reaction) are shown in Figure 1. The modal response for engagement, satisfaction and relevance of the session was ‘strongly agree’ with over 80% of students opting for this response.

**Figure 1:**
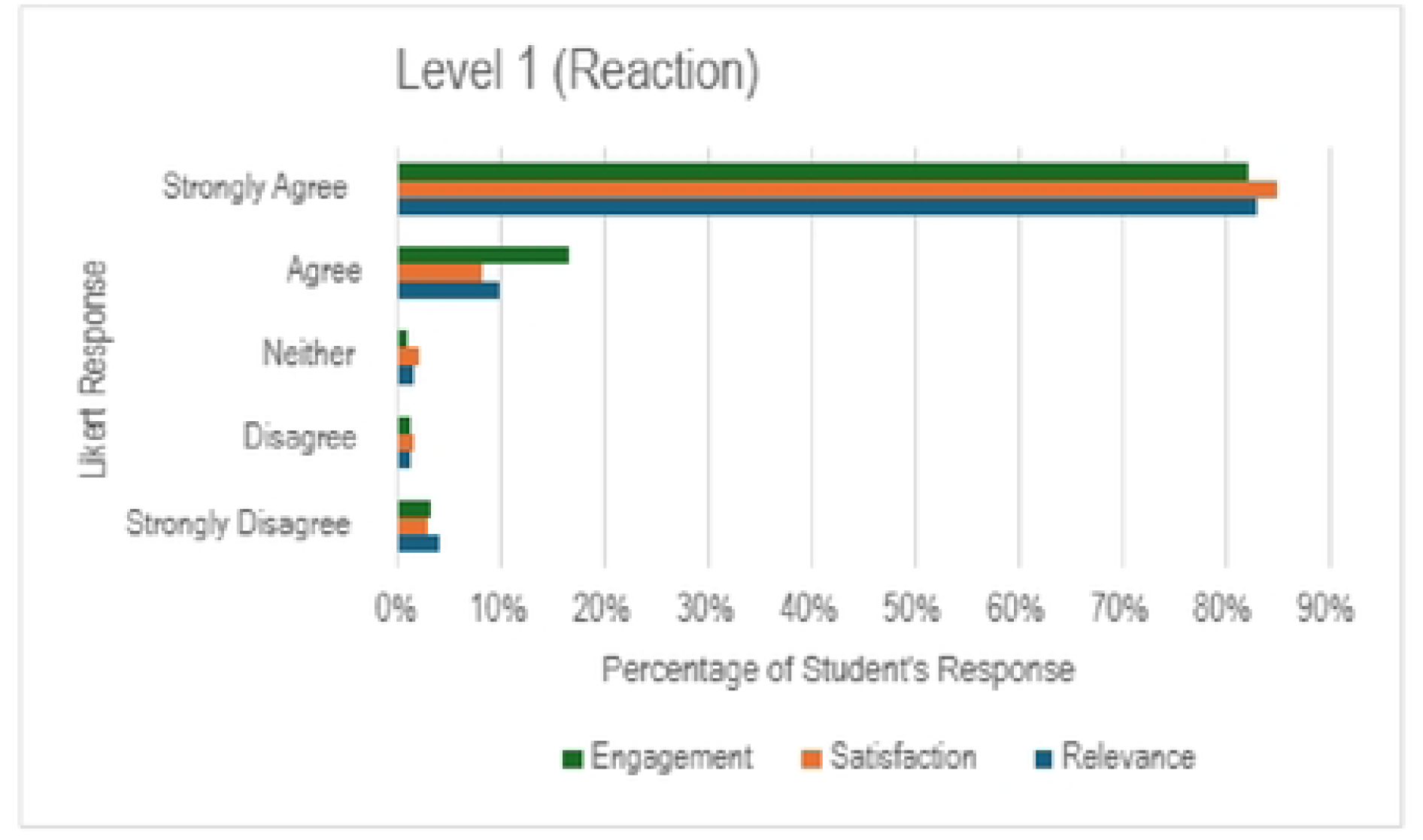
Frequency of responses for Level 1 (Reaction)

When considering Learning (Figure 2), strongly agree was again the modal response in all areas (Skills & Knowledge, Attitude, Confidence and Commitment). With the exception of ‘Confidence’, 76% and above of students opted for ‘strongly agree’. While confidence generated a response of strongly agree for 60% of students, it is worth noting that agree and strongly agree responses combined accounted for over 80% of student’s responses.

**Figure 2:**
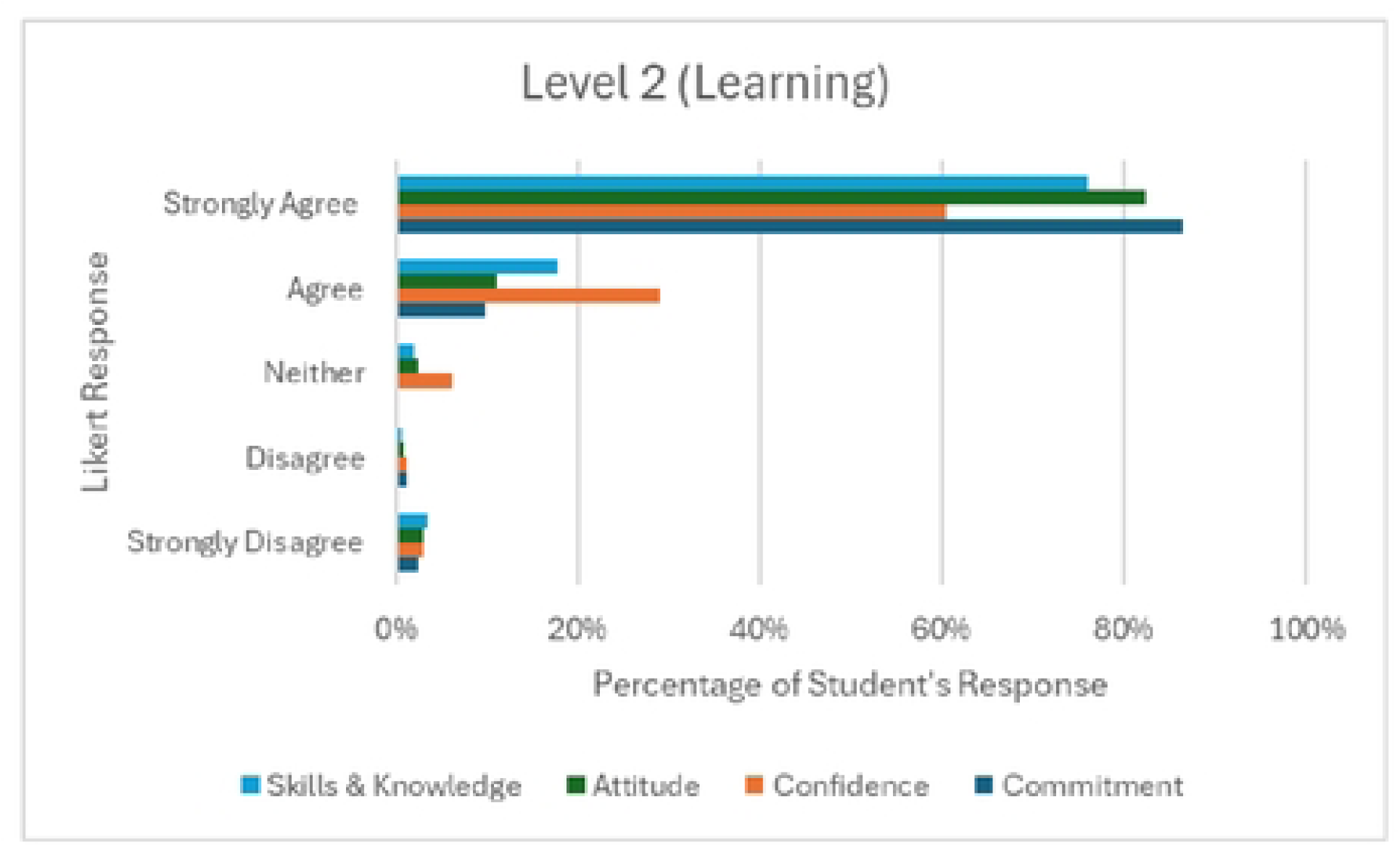
Frequency of responses for Level 2 (Learning)

The focus groups generated 16 codes which were then grouped into 6 themes: (1) Reaction, (2) Learning, (3) Participation, (4) Placement, (5) Skills and (6) Suggestions (Table 2).

**Table 2:** Focus Group Themes and Codes.

| Theme | Codes | Illustrative Quotes |
| --- | --- | --- |
| Reaction | -Likes<br>-Dislikes<br>-Feelings pre SBE | It let you see how you would react. (P7)*<br>I just hate when people are watching me. (P1)<br>There was a bit of nervousness and anxiousness. (P6) |
| Learning | -Comparison to other teaching methods<br>-Experience | You can take the benefit from SBE as you've done the theory. (P1)<br>The most helpful part was the debrief after. (P4)<br>With SBE, we all got a chance of taking part, we all got feedback. (P6) |
| Participation | -Exposure<br>-Timing<br>-Frequency | I haven't seen an acute patient...I wasn't even used to the set-up of a hospital bed. (P1)<br>You go in blind, like in your first year....if you do have SBE at least you will....be more confident. (P2)<br>There really wasn't enough of it (*SBE throughout the programme*). (P1) |
| Placement | -SBE in Preparation for Placement<br>-SBE as a Substitute for Placement | It's (*SBE*) good for placement and on call and stuff because you're actually doing it. (P2)<br>I wouldn't say it would be a substitute for it (placement).... I find working on real patients a lot different even to SBE, to be honest. I think placement has so many benefits. (P8)<br>I don't see why it wouldn't be classed as placement hours. (P1)<br>I would feel quite nervous if it was replacing placement hours. I'd rather just add on them if that makes sense. (P4) |
| Skills | -Adaptation<br>-Communication<br>-Confidence<br>-Decision Making<br>-Standing Back<br>-Teamwork | You had to make decisions quickly... I don't fully trust my decisions. (P1)<br>You work up in your head what you will do .... Then you have to think on your feet. (P3)<br>It (*SBE*) teaches you to stand back and take in the whole situation. (P6) |
| Suggestions |  | To alleviate some nerves.... show students a video of what you kind of expect from like a previous year group or something. (P8)<br>Actors would be interesting to use. (P5)<br>Maybe making it (SBE) neuro based...the assessment in neuro is so so long....nice to rehearse. (P5) |

*\*P & Number = Participant*

### Reaction

Students reported being worried prior to the session largely around the fear of being ‘wrong’ in front of peers. While they liked the hands-on experience and immediate feedback, they did not like that others were watching them.

*So the fear wasn’t long going - It was more the fear of the unknown until you got into the situation.* (P6)

### Learning

Students ranked SBE highly in comparison to other learning and teaching strategies. They did note the benefit of having underpinning theory from previous lectures/practical’s which made the SBE session more effective. Earning came from the ‘real world’ feel of case studies and the debrief.

*The bit that I find most helpful during the actual simulation part was the debrief that we had after it. We just talked quite informally*. (P4)

### Participation

Those students who hadn’t had the opportunity to experience an acute environment or patient, welcomed the chance of exposure and felt it put them on a more equal footing with those who had. Consensus was that students wanted more SBE and also earlier in their physiotherapy programme.

*It sets everybody on an equal playing field. Some people here have had really good placement in ICU and have been really hands on, and other people haven’t. I think it would make everybody on an equal playing field for what they know and the exposure they have to it.* (P4)

### Placement

Students felt that SBE was good preparation for placement and on-call duties. While they viewed SBE as a useful supplement to clinical placement perhaps contributing to hours, they were not keen that SBE should fully substitute placement.

*I think if you’re really stuck for hours, yeah, it could be used as an alternative, but I think there’s really important aspects of being in a hospital.* (P5)

### Skills

Students highlighted transferable skills gained by SBE namely standing back to taking in the whole situation and adapting treatment direction where necessary. Other themes identified were the need for good communication and teamwork. Students also noted an improvement in confidence given the necessity to engage in decision making.

*It gives you that environment where you can be forced to make decisions. To me that’s a good skill to develop because when you go to work, nobody’s going to make the decisions for you.* (P1)

*So, it kind of prepared you - you were just able to take a step back, think about it before you would decide on what to do*. (P6)

### Suggestions

Students suggested prior to SBE having an information video from previous students who participated by way of helping them prepare for what to expect. Some students would welcome actors as patients. Neurology was highlighted as an area that students would welcome SBE scenarios.

*I haven’t done a neuro placement. Obviously, we’ve only got one left, so if I do get neuro, I’m not going to lie, I am really panicking about doing it. I feel like I really would have benefited if I had something similar to what we did from a neuro perspective*. (P1)

## DISCUSSION

The qualitative and quantitative data collected in this SBE study outlined a positive student learning experience, similar to findings from other physiotherapy students [15]. A recent scoping review acknowledged the growing body of evidence in SBE within physiotherapy, however pointed out that much of this work has been undertaken within the cardiorespiratory field [16]. These authors suggested further exploration of SBE in other areas of physiotherapy. This is in keeping with the qualitative feedback in this study, where students indicated that they would welcome more SBE, both earlier in their programme and across different clinical areas, namely neurology. It is important however to be mindful of the holistic approach endorsed by the Chartered Society of Physiotherapy (CSP) Framework [17], perhaps taking less of a siloed systems approach, but instead adopting a person centred one. Irrespective of the clinical area of content, student feedback in this study did highlight attainment of transferable skills such as communication, decision making, adaptability and teamworking which they took forward to clinical placement.

Students also voiced that SBE helped to ‘level the playing field’ in terms of opportunity with exposure to critical care that not everyone will get the chance to experience on clinical placement. This is an approach that was adopted in a paediatric study with an outcome of improved student self-efficacy [18]. The positivity towards SBE in this study is consistent with existing research. SBE was found to be effective in physiotherapy education and improved students’ learning experience [19]. The reasons to explain this however are complicated give the complex learning processes in healthcare and the difficulty linking theory and practice is a continued challenge [20]. Higher education curriculum design should strive to embody active learning, inclusivity and employability, all of which SBE stives to do in preparation for working life [21]. Incorporation of these elements could be part of the appeal for students. Another reason could be the underpinning pedagogical design of SBE, although SBE has been described not as a pedagogy but rather an immersive learning platform incorporating three design principles [22]. These principles are Behaviourist (rote learning and repetition), Cognitivism (active participant including observation) and Constructivism (new habit formation through experiences). The classroom may be too controlled or unrealistic to achieve these principles, and the clinical environment may be too uncertain or risky. SBE therefore provides a ‘sweet spot’. In the focus groups, students rated SBE above other forms of learning and teaching. This notion was echoed in a medical education review [23]. Despite concerns about financial investment and maintaining realism, this medical review noted the benefit SBE offers a safe and controlled environment to hone technical and non-technical skills, ultimately improving patient safety. The effectiveness of SBE was underscored as being above that of traditional teaching methods [23].

The concept of emergence in SBE embraces the notion that (despite these three design principles), stability and change can co-occur within SBE [24]. In other words what unfolds may not be what was planned. Students therefore get the opportunity to react and make mistakes in that ‘safe space’. This also feeds into the notion of employability and feeling practiced and ready for the unpredictable real world. It should be noted however that the concept of a safe space and psychological safety does not just happen but needs to be created and nurtured, and the metaphor of ‘tending the garden’ to reap rewards could be used in the pursuit of psychological safety [25].

Other professions such as nursing have noted similar results as this study, noting an increase in confidence, teamwork and communication [26]. In this study however, the confidence domain in level 2 (learning) was the area where students were less positive in comparison to other areas. During the focus group sessions students also highlighted a lack of confidence prior to commencing the SBE session. One way of mitigating this may have been to utilise co-production of simulation scenarios with students to add to the learning experience [27]. Further exploration of confidence pre and post SBE session as outlined in the “New World” Kirkpatrick Level 2 outcomes [28] may have been beneficial. It is also important to note that whilst time was spent ensuring the psychological safety of learners [29] the students had no real experience of SBE prior to this session and earlier exposure in the degree programme might enhance student confidence. This point was supported in some of the qualitative feedback during the focus groups sessions with students stating they would like earlier and more exposure to SBE along with materials to support the process.

The lower scores in confidence may also be rooted in the students’ inability to ‘suspend their disbelief.’ This is the ability to believe the unbelievable and accept lapses in authenticity in order to fully immerse in the experience [30]. It is accepted that high fidelity SBE does not necessarily equate to high quality SBE [31]. Thinking beyond fidelity, planning SBE sessions must focus however on a sense of realism. In the scenarios in this study, manikins were utilised (it would not be feasible, safe or ethical to use an actual ventilated patient). It may be hard for students to ignore that fact, and they may not feel truly confident until they have undertaken the treatment on a real patient.

The idea of still needing ‘practice’ on real patients was reinforced during the qualitative feedback within the focus group sessions. Students viewed SBE as a useful bridge between the classroom and the clinical environment. They felt SBE could be a useful addition to clinical placement, perhaps used to supplement hours but not fully replace the placement experience. Undoubtedly pressures exist in accessing clinical placements for all physiotherapy students. This is illustrated on the island of Ireland, where growth in programmes threaten traditional 1:1 models of supervision [32]. A study involving nurses and allied health professionals reported that simulation could replace 11-30% of traditional practice placement time however it is important to note that only 3 of the participants were physiotherapists [33]. Research has shown also that switching up to 25% of practice learning to a simulated model does not compromise student attainment or competency [34]. Specifically in physiotherapy a small pilot study which replaced the first week of clinical placement with SBE showed that confidence to undertake placement was improved, and attainment of competence unaffected by the reduction in clinical hours [35]. Within the cardiorespiratory area of physiotherapy, a hybrid approach combining SBE with clinical hours (as opposed to replacing them) showed that clinical competency requirements were satisfied [36]. Whilst adoption of this hybrid model may also be advocated by students in this study, significant barriers remain which threaten success. Realism continues to be a key facilitator of a successful simulated placement and student exposure [27]. Funding, training, logistics and meaningful engagement with all stakeholders is required [32]. This process should be in parallel with a cost/value analysis, to prove the worth of SBE in terms of viability and sustainability [37]. Modelling of costs is not exact or standardised and should consider not simply monetary but social benefits. Research has demonstrated a significantly positive cost benefit ratio [38]. Given the logistics involved in planning, it is unlikely that SBE would be a quick fix to placement issues but rather a longer-term strategy, which would require vision. Often (but not always) in modelling costs are concentrated in the early phase with benefits not reaped until much later [38]. This could be off putting for policy makers who want to see a quick reward for their investment.

The findings of this study are consistent with other research in terms of the benefit SBE can bring to students. In the pursuit of sustainable, high quality SBE the global consensus Call for Action must be remembered whereby policy makers are urged to embrace and support the development of SBE [39].

## CONCLUSION

In keeping with existing research, students in this study rated SBE positively with calls for more opportunities, earlier in the programme and spanning clinical areas beyond cardiorespiratory. Students also felt SBE was a good way to ‘level the playing field’ and gain exposure to clinical areas they may not get the opportunity to be exposed to on placement. Non-technical skills were developed which were transferred into the clinical environment, irrespective of the clinical area. Reaction to SBE was high in terms of relevance, engagement and satisfaction. Students rated it higher than other forms of learning and teaching. When considering learning achieved, confidence although positive, was the relatively lowest scoring domain. This could perhaps be attributed to the lack of realism, leading to the inability to suspend disbelief.

Students expressed that while they saw huge value in SBE, they would be wary of it totally replacing clinical placement but advocated a more hybrid approach. Adoption of such a model may be welcome given the pressures on placing students, but would require much vision, investment, standardisation, planning and cost/benefit analysis. This may make it a longer-term strategy requiring support of policy makers.

## Data Availability

The minimal data set is available at PhysioNet via https://doi.org/10.13026/*****

## ETHICAL APPROVAL

Ethical approval was granted by the Ulster University Nursing and Health Sciences Filter committee (Reference: FCNUR-24-072).

## ACKNOWLEDGEMENTS

The authors wish to thank the final year cohort of physiotherapy students (Academic year 2024/25) for their participation and valuable contributions.

## REFERENCES

1. Hawker C, Jones B, Cooke S, Mitra S, Hoole A, Bartholomew B et al. Developing an all-wales definition of simulation-based education:. International Journal of Healthcare Simulation. 2022;2.(Supplement 1):A40–41 10.54531/INHM4618

2. Maran NJ, Glavin RJ. Low- to high-fidelity simulation – a continuum of medical education? Medical Education 2003;22–8.

3. Thomas EM, Rybski MF, Apke TL, Kegelmeyer DA, Kloos AD. An acute interprofessional simulation experience for occupational and physical therapy students: Key findings from a survey study. Journal of Interprofessional Care. 2017;31:317–324. doi:10.1080/13561820.2017.1280006

4. Berg B. Simulation: past, present, and future. Clin Exp Emerg Med.2023;10(2):125–128. doi: 10.15441/ceem.23.044.

5. Komasawa N, Yokohira M. Simulation-based education in the artificial intelligence era. Cureus. 2023;15(6).

6. Heneghan N, Thackery D, Stiger R, Aliner G, Jennings J, Spearpoint K et al. Simulation toolkit for pre-registration physiotherapy education/training. 2023; (PDF)Simulation toolkit for pre-registration physiotherapy education/training

7. Minns Lowe C, Heneghan N, Herbland A, Atkinson K, Beeton K. KNOWBEST: The KNOWledge, BEhaviours and Skills required of the modern physioTherapy graduate including the future role of practice-based learning. CSP. 2022.

8. Yardley S, Dornan S. Kirkpatrick’s levels and education ‘evidence’. Medical Education. 2012;46:97–106. 10.1111/j.1365-2923.2011.04076.x

9. Mansell SK, Grafton K, Barnfield E, Eckersley G, Bendall A, Cork G et al. Simulation-based education within respiratory physiotherapy training: a scoping review. ACPRC Journal. 2024;37–51. doi:10.56792/KEPM1936

10. Squires K, Heaney S, MacDonald-Wicks L, Johnston C, Brown L. Mapping Simulated-Based Learning Experiences Incorporated into Professional Placements in Allied Health Programs: A Scoping Review. Simulation in Healthcare: The Journal of the Society for Simulation in Healthcare. 2022;17(6), 403–415. DOI: 10.1097/SIH.0000000000000627

11. Johnston S, Coyer FM, Nash R. Kirkpatrick’s Evaluation of Simulation and Debriefing in Health Care Education: A Systematic Review. Journal of Nursing Education. 2018;1;57(7):393–398. doi: 10.3928/01484834-20180618-03.

12. Dairo YM, Hunter K, Ishaku T. The impact of simulation-based learning on the knowledge, attitude and performance of physiotherapy students on practice placements. BMC Medical Education. 2024; 24(1):786. doi: 10.1186/s12909-024-05718-2.

13. Levett-Jones T, McCoy M, Lapkin S, Noble D, Hoffman K, Dempsey J et al. The development and psychometric testing of the Satisfaction with Simulation Experience scale. Nurse Education Today. 2011; 31(7):705–10. doi:10.1016/j.nedt.2011.01.004

14. Braun V, Clarke V. Using thematic analysis in psychology. Qualitative Research in Psychology. 2006;3, 77–101.

15. Mori B, Carnahan H, Herold J. Use of Simulation Learning Experiences in Physical Therapy Entry-to-Practice Curricula: A Systematic Review. Physiother Can. 2015;67(2):194–202. doi: 10.3138/ptc.2014-40E.

16. Room J, Stiger R. The use of simulated learning in prequalifying physiotherapy education: a scoping review. Int J of Healthcare Simulation. 2022; 3(1):A16–A16 doi:10.54531/HVPN9537,

17. CSP. Physiotherapy Framework: putting physiotherapy behaviours, values, knowledge & skills into practice [updated May 2020] 2011;csp_physiotherapy_framework_0.pdf

18. Hough J, Levan D, Steele M, Kelly K, Dalton M. Simulation-based education improves student self-efficacy in physiotherapy assessment and management of paediatric patients. BMC Med Educ.2019;16;19(1):463. doi: 10.1186/s12909-019-1894-2.

19. Koukourikos K, Tsaloglidou A, Kourkouta L, Papathanasiou IV, Iliadis C, Fratzana A et al. Simulation in Clinical Nursing Education. Acta Inform Med. 2021;29(1):15–20

20. Valaitham J. A systematic review of the effectiveness of Simulation in Physiotherapy Education. 2023;[A SYSTEMATIC REVIEW OF THE EFFECTIVENESS OF SIMULATION IN PHYSIOTHERAPY EDUCATION | World Physiotherapy](https://world.physio/congress-proceeding/systematic-review-effectiveness-simulation-physiotherapy-education-0)

21. Murphy C, Curran R. Empowering curriculum leaders to innovate: An overview of an Integrated Curriculum Design Framework. Educational Developments, SEDA Ltd. 2020;Issue 21.1:17–21.

22. Gwen D, Smythe L, Wright-St Clair V. Simulation is not a pedagogy. Open J of Nursing. 2017;7(7):779 doi 10.4236/ojn.2017.77059

23. Elendu C, Amaechi D, Okatta A, Amaechi E, Elendu T, Ezeh C et al. The impact of simulation-based training in medical education: A review. Medicine 2024;103(27):e38813 DOI: 10.1097/MD.0000000000038813

24. Rooney D, Hopwood N, Boud D, Kelly M. The role of simulation in pedagogies of higher education for the health professions: Through a practice-based lens. Vocations and Learning, 2015;8(3), pp.269–285.

25. Nestel D, Bruun B, Dieckmann P, Tulloch S, Gormley G. ‘Tending’ the ‘garden’ of psychological safety in simulation-based education. Journal of Healthcare Simulation. 2025 from 10.54531/CPZH5763.

26. Alrashidi N, Pasay an E, Alrashedi MS et al. Effects of simulation in improving the self-confidence of student nurses in clinical practice: a systematic review. BMC Med Educ 2023;23:815. 10.1186/s12909-023-04793-1

27. Stevens K, Sathe K, Mathew C, McLean S. Experiences of students and educators with simulated placements in allied health profession and nursing education: a qualitative systematic review:. International Journal of Healthcare Simulation. 2023; from 10.54531/ftwz5026.

28. Kirkpatrick J. An Introduction to the New World Kirkpatrick Model. Kirkpatrick Partners; 2015:2019. doi:10.1515/9781580468619

29. Gaba D. Simulations That Are Challenging to the Psyche of Participants: How Much Should We Worry and About What?. Simulation in Healthcare: The Journal of the Society for Simulation in Healthcare. 2013;8(1): 4–7DOI: 10.1097/SIH.0b013e3182845a6f

30. Muckler V. Exploring Suspension of Disbelief During Simulation-Based Learning. Clinical Simulation In Nursing. 2017; 13(1): 3 – 9

31. Richardson C, Thompson J, Jacklin S. An orange will do: Suspending learner disbelief in simulations. Currents in Pharmacy Teaching and Learning. 2022;14(11):1337–1339 10.1016/j.cptl.2022.09.016.

32. O’Shea O, Mulhall C, Condron C, McDonough S, Larkin J, Eppich W. A qualitative study of physiotherapy educators’ views and experience of practice education and simulation-based learning:. International Journal of Healthcare Simulation. 2023; from 10.54531/hkoi8650.

33. Bridge P, Adeoye J, Edge C, Garner V, Humphreys A, Ketterer S, et al. Simulated placements as partial replacement of clinical training time: a Delphi consensus study. Clinical Simulation in Nursing. 2022;68:42–8

34. Wright A, Moss P, Dennis D, Harrold M, Levy S, Furness A et al. The influence of a full-time, immersive simulation-based clinical placement on physiotherapy student confidence during the transition to clinical practice. Advances in simulation. 2018;3(1):1–0.

35. Blackford J, McAllister L, Alison J. Simulated Learning in the Clinical Education of Novice Physiotherapy Students. International Journal of Practice-Based Learning in Health and Social Care. 2015; 3(1), 77–93. 10.18552/ijpblhsc.v3i1.209

36. Blackstock F, Watson K, Morris N. Simulation Can Contribute a Part of Cardiorespiratory Physiotherapy Clinical Education: Two Randomized Trials. Simulation in Healthcare: The Journal of the Society for Simulation in Healthcare. 2013; 8(1):32–42. DOI: 10.1097/SIH.0b013e318273101a

37. Maloney S, Haines T. Issues of cost-benefit and cost-effectiveness for simulation in health professions education. Adv Simul (Lond). 2016;17(1)13. doi: 10.1186/s41077-016-0020-3. PMID: 29449982; PMCID: PMC5806357.

38. Soorapanth S, Eldabi T, Young T. Towards a framework for evaluating the costs and benefits of simulation modelling in healthcare. The Journal of the Operational Research Society. 2023;74(3):637–646 DOI: 10.1080/01605682.2022.2064780

39. Diaz-Navarro C, Armstrong R, Charnetski M, Freeman K, Koh S, Reedy G et al. Global consensus statement on simulation-based practice in healthcare. Adv Simul. 2024;9, 19 10.1186/s41077-024-00288-1

